# Prediction of gestational diabetes mellitus using early oral glucose tolerance test

**DOI:** 10.1101/2022.03.29.22273154

**Authors:** C. Kandauda, S.S Manathunga, I.A Abeyagunawardena, K.M.H.C Thilakarathne

## Abstract

**Introduction:** Gestational Diabetes Mellitus (GDM) is defined as diabetes first detected at the second or third trimester of pregnancy, excluding preexisting diabetes. We aimed to build a predictive model of GDM using booking oral glucose tolerance test (OGTT) values.

**Materials and Methods:** Seventy-five healthy mothers who underwent 75g OGTT at 12-14 weeks and at 24-28 weeks were recruited. GDM was diagnosed at 28 weeks by cutoffs proposed by the Hyperglycemia and Adverse Pregnancy Outcomes study.

Sensitivities and specificities for diagnosing GDM using different cut-offs for each of the three booking OGTT variables were measured. A series of multivariate binary logistic regression models were fitted using different combinations of the three booking OGTT variables. In-sample sensitivities and specificities for different cutoff probabilities of the models were calculated and Receiver Operating Characteristic (ROC) curves were constructed. The Area Under the Curve (AUC) of the ROC curve and the best cutoff value which maximized the sum of sensitivity and specificity of each model were computed.

**Results:** AUC of ROC curves for isolated fasting, 1 hour and 2 hour booking OGTT values for the prediction of GDM were 69.8%, 67.1% and 61.0% respectively. However, the logistic regression model with fasting and 1 hour booking OGTT values as predictors out-performed all other models with an AUC of 76.3%, in-sample sensitivity of 87.5% and a negative predictive value of 95.12%.

**Conclusions:** The future occurrence of GDM can be predicted utilizing a logistic model with fasting and 1 hour booking OGTT variables, which enables early identification and intervention.

## Introduction

Gestational diabetes mellitus (GDM) remains one of the most common medical complications of pregnancy and is associated with an array of maternal and fetal adverse outcomes. It is defined as glucose intolerance with onset or first recognition in pregnancy [1]. GDM can result in macrosomia, shoulder dystocia, preterm delivery, preeclampsia, neonatal hypoglycemia, neonatal hyperbilirubinemia leading to prolonged hospital stays. Hence, meticulous screening programs are implemented worldwide to screen for GDM with the aim of taking preemptive actions to control hyperglycemia in pregnancy thus reducing adverse outcomes [2].

There had been controversies on the diagnosis and screening of GDM [3]. The 2002 National Institute for Health and Care Excellence (NICE) Health Technology Assessment concluded that there was insufficient evidence to advocate universal screening in pregnancy. Therefore, the 2008 NICE guidelines recommends pregnant women with BMI above 30 kg/m2, a previous macrosomic baby weighing 4.5 kg or more, previous gestational diabetes, a family history of diabetes (first-degree relative with diabetes) or belonging to an ethnicity with a high prevalence of diabetes identified during the booking visit to be offered screening with the two hour oral glucose tolerance test (OGTT) [4].

GDM is diagnosed if the fasting blood glucose value is more than/equal to 5.1mmol/l (92 mg/dl), 1 hour plasma glucose more than/equal to 10.0 mmol/l (180 mg/dl) or if 2 hour blood glucose is more than/equal to 8.5mmol/l (153 mg/dl) according to the hyperglycemia associated pregnancy outcomes (HAPO) study [2]. Women with no risk factors undergo OGTT if glycosuria more than 2+ is detected in the routine urinary dipstick testing. For patients with a history of GDM, OGTT is performed as soon as possible after the booking visit and at 24-28 weeks if the initial test is normal. For women with other risk factors OGTT is performed at 24-28 weeks to detect GDM [4].

This study aimed to build a predictive model for occurrence of GDM in high-risk women with normal OGTT values at booking visit.

## Materials and Methods

The study was conducted at the antenatal clinics in Kandy, Sri Lanka. Ethical clearance was obtained from the Ethics Review Committee, Faculty of Medicine, University of Peradeniya. A total of 75 mothers with no other comorbidities, who had undergone a 75g OGTT at booking visit between 12-14 weeks and at 24-28 weeks were recruited. Written informed consent was taken from all subjects. Diagnosis of GDM at 28 weeks was made based on Hyperglycemia and Adverse Pregnancy Outcomes (HAPO) study cutoffs [5].

The distribution of fasting plasma glucose values, one-hour plasma glucose values and two-hour plasma glucose values were visualized using density curves for the GDM and non-GDM samples. Correlations between the booking and the corresponding 28-week plasma glucose values were calculated and separate Receiver Operating Characteristic (ROC) curves were constructed for fasting, one-hour and two-hour booking plasma glucose values, for the diagnosis of GDM.

A series of multivariate binary logistic regression models were fitted using different combinations of booking OGTT predictors for the prediction of GDM. In-sample sensitivities and specificities for different cutoffs of log of odds ratios of the logistic models were calculated. ROC curves were constructed and the Area Under the Curve (AUC) of the ROC curve and the best cutoff value which maximized the sum of sensitivity and specificity of each model were computed.

## Results

Distribution of the fasting, one-hour and two-hour booking plasma glucose values are visualized in Fig 1, Fig 2 and Fig 3 respectively.

**Fig 1.**
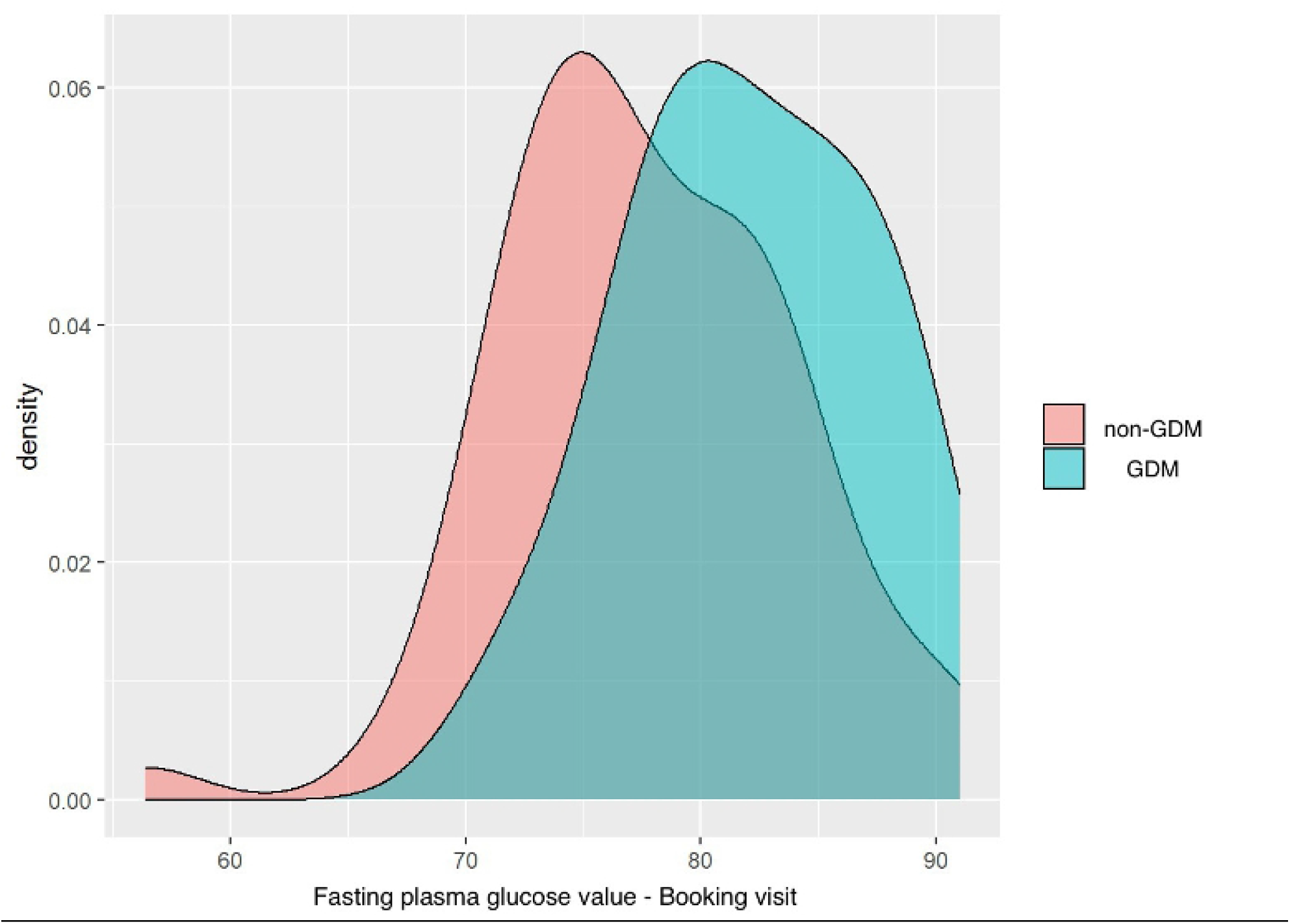
Distribution of the fasting blood glucose values in GDM and non-GDM populations at the booking visit

**Fig 2.**
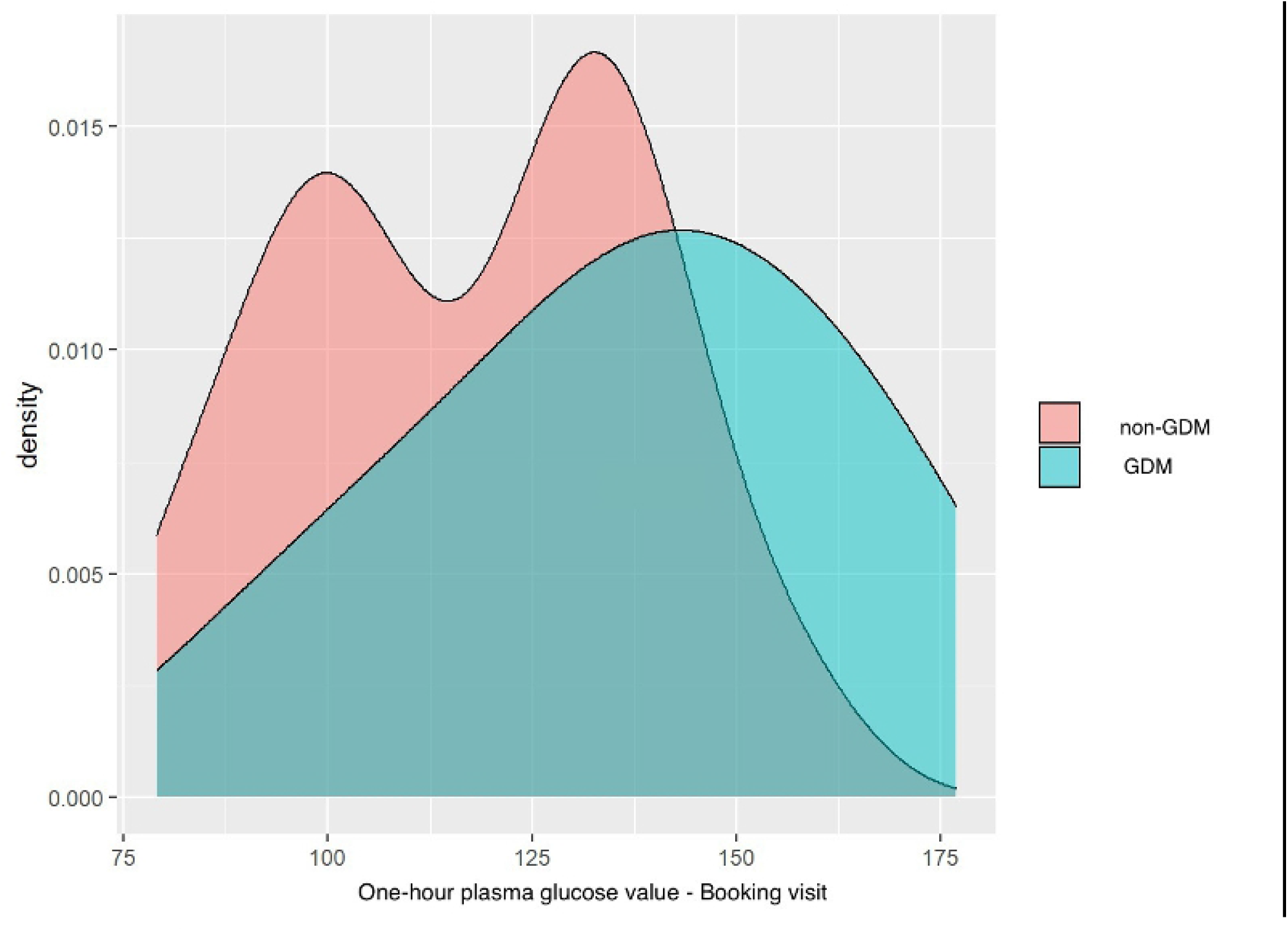
Distribution of the one-hour blood glucose values in GDM and non-GDM populations at booking visit

**Fig 3.**
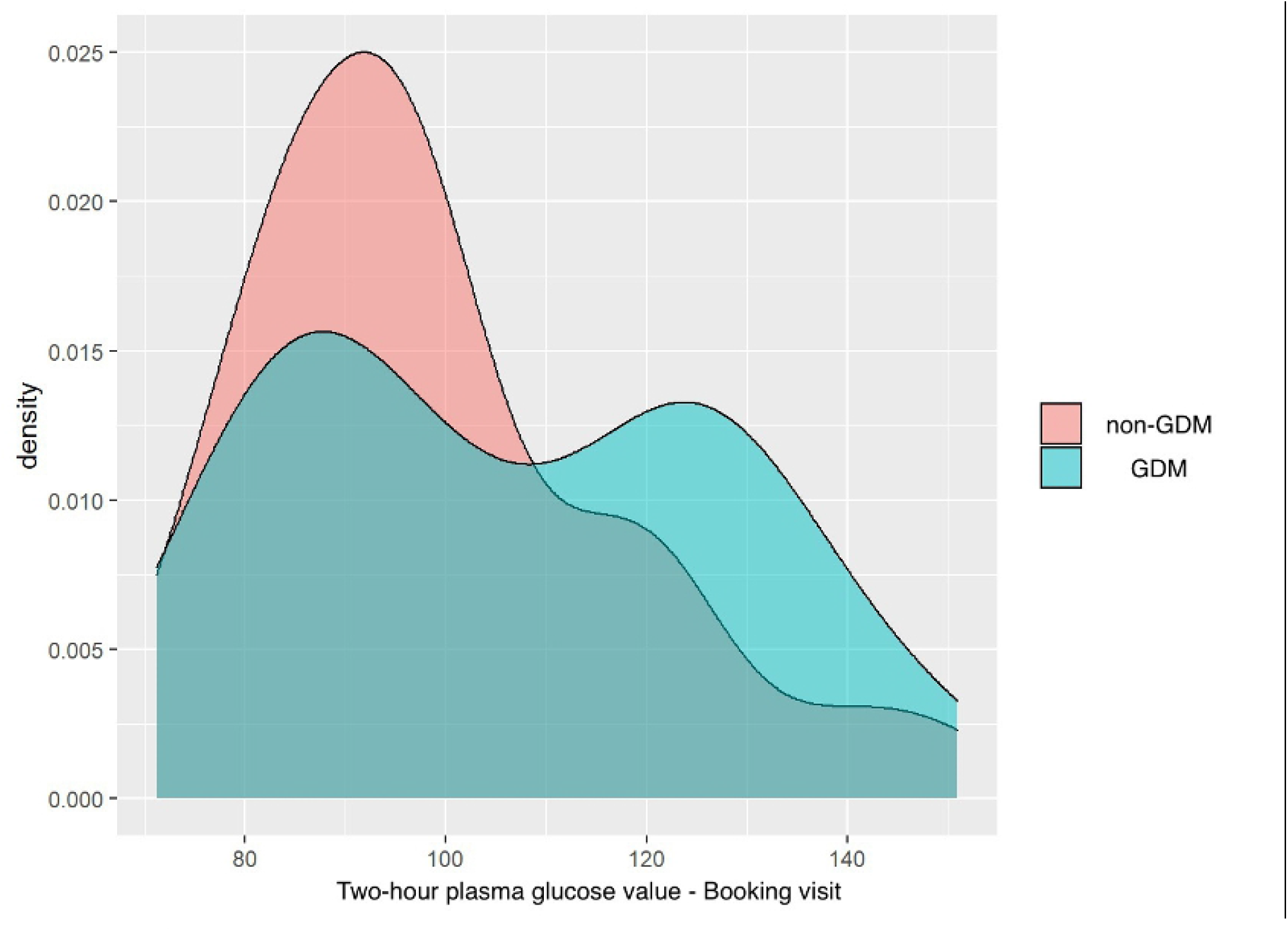
Distribution of the two-hour blood glucose values in GDM and non-GDM populations at booking visit

It can be observed that the plasma glucose values are shifted rightwards in the GDM samples when compared to non-GDM samples. The correlation coefficients of fasting, one-hour and two-hour plasma glucose values between the booking and 28-weeks tests of GDM and non-GDM mothers are 0.29, 0.58 and 0.36 respectively as depicted in Fig 4, Fig 5 and Fig 6 respectively. The Pearson Correlation coefficient was utilized.

**Fig 4.**
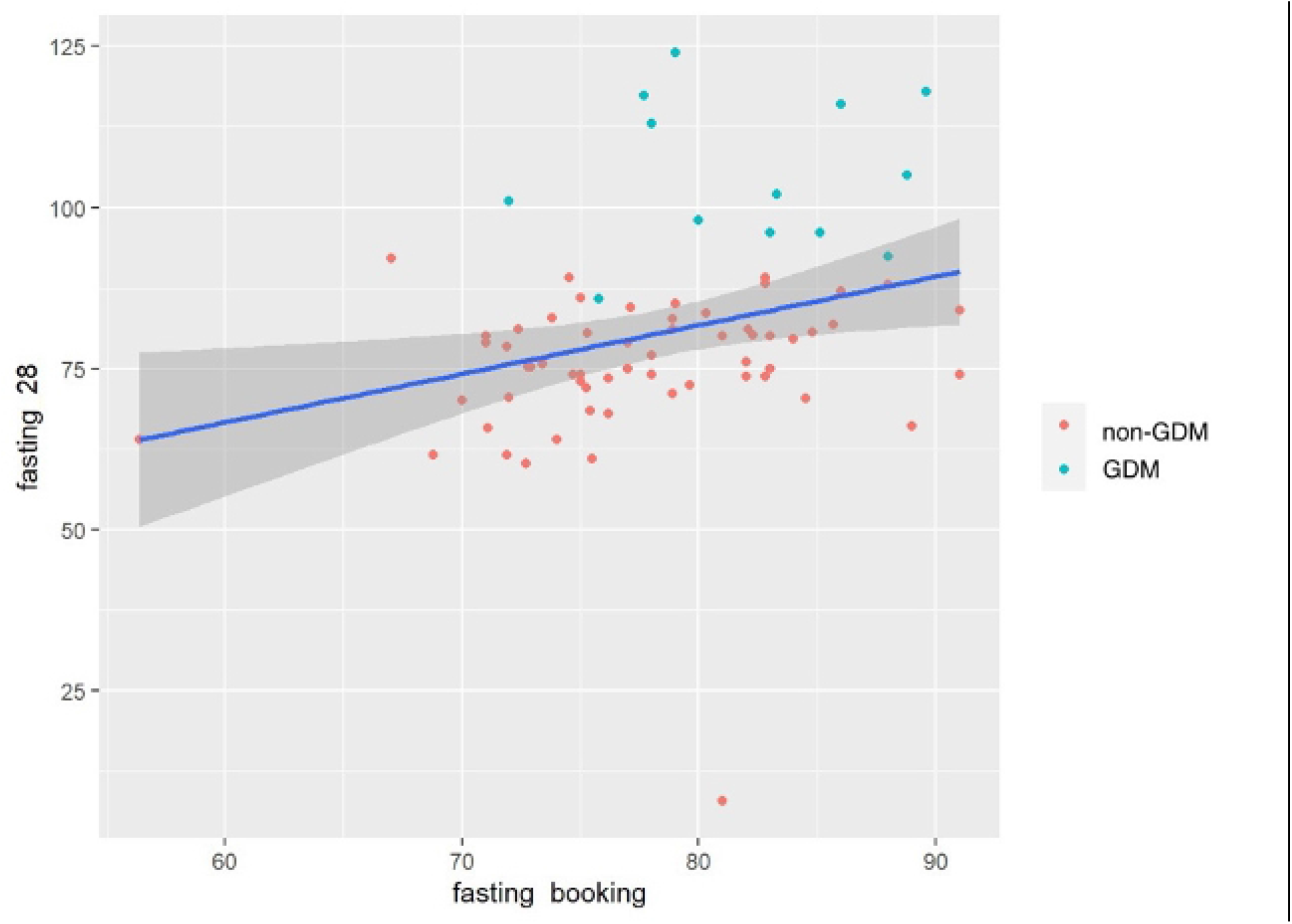
Scatterplot depicting correlation coefficient of fasting blood glucose value of GDM and non-GDM mothers between booking visit and 28 weeks

**Fig 5.**
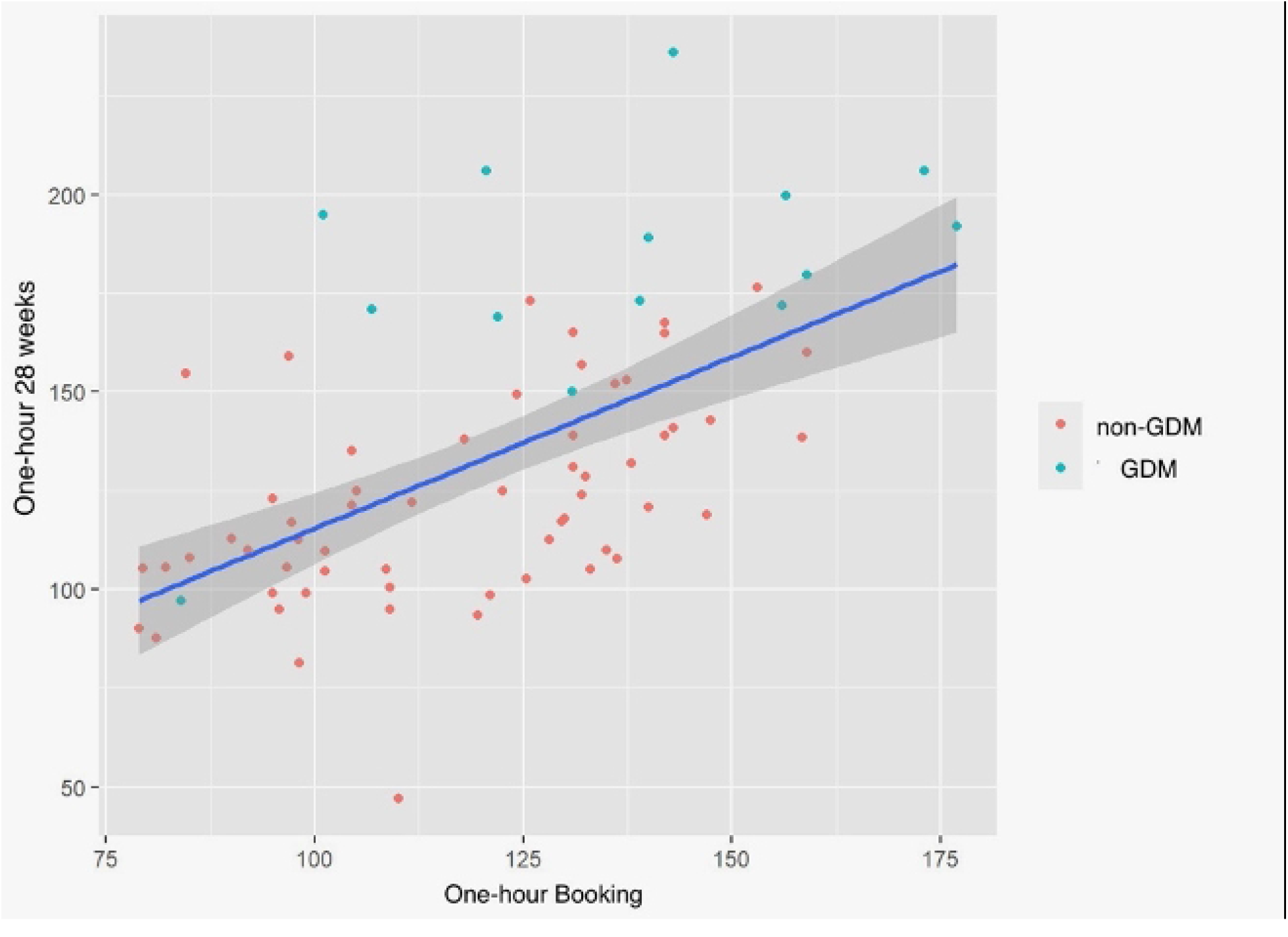
Scatterplot depicting correlation coefficient of one-hour blood glucose value of GDM and non-GDM mothers between booking visit and 28 weeks

**Fig 6.**
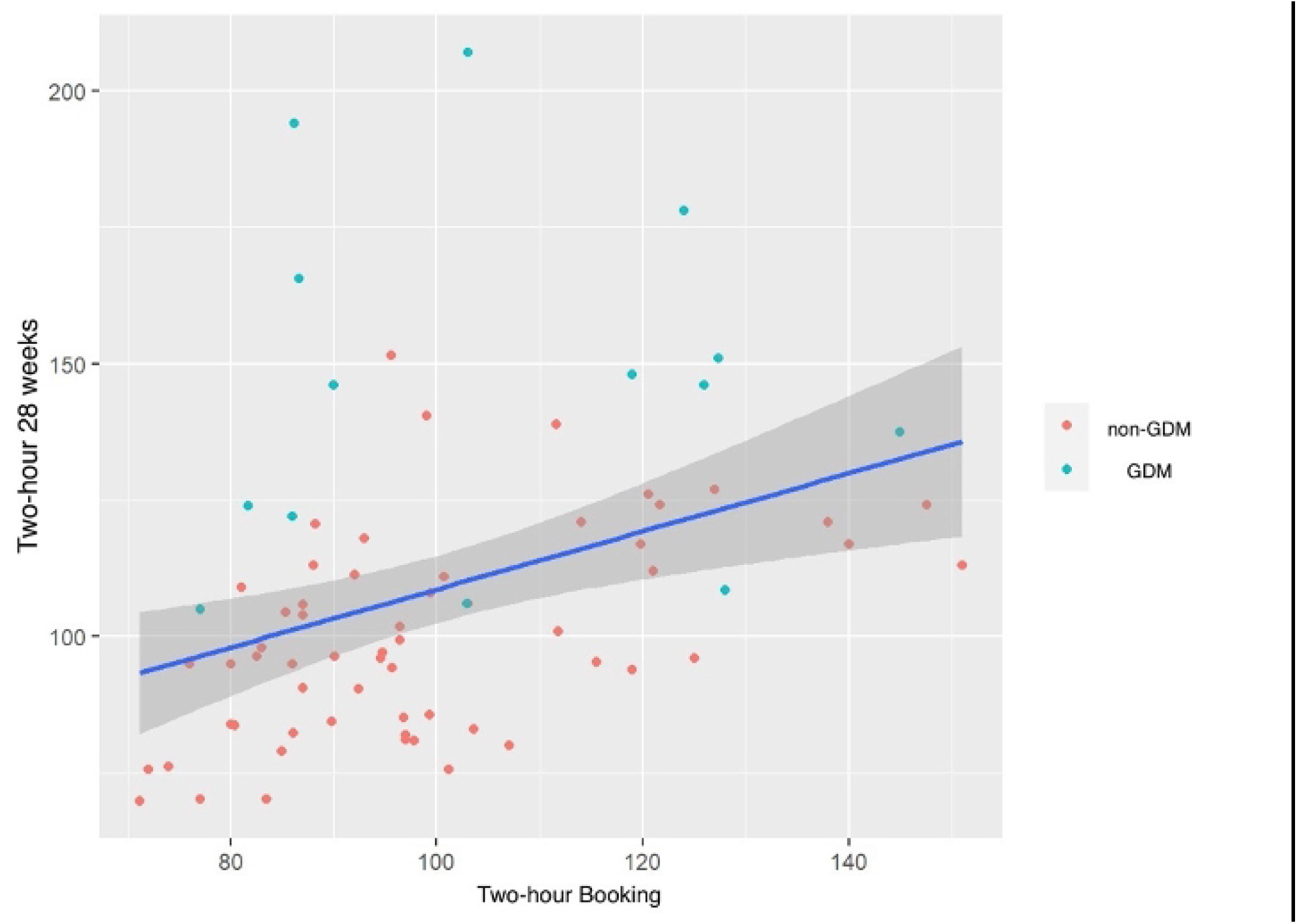
Scatterplot depicting correlation coefficient of two hour blood glucose value of GDM and non-GDM mothers between booking visit and 28 weeks

The discriminative powers of the isolated booking plasma glucose values are summarized in the following Table 01.

**Table 01.** Discriminatory powers of individual booking visit plasma glucose values

| Variable | Cutoff value<br>(mg/dl) | Sensitivity (%) | Specificity (%) | AUC (%) |
| --- | --- | --- | --- | --- |
| Fasting plasma<br>glucose | 77.4 | 85.7 | 52.5 | 71.2 |
| One-hour<br>plasma glucose | 138.5 | 57.1 | 83.1 | 70.4 |
| Two-hour<br>plasma glucose | 102.1 | 57.1 | 69.5 | 59.0 |

The ROC curves for individual booking visit plasma glucose values for fasting, one hour and two hours are illustrated in Fig 7, Fig 8 and Fig 9 respectively.

**Fig 7.**
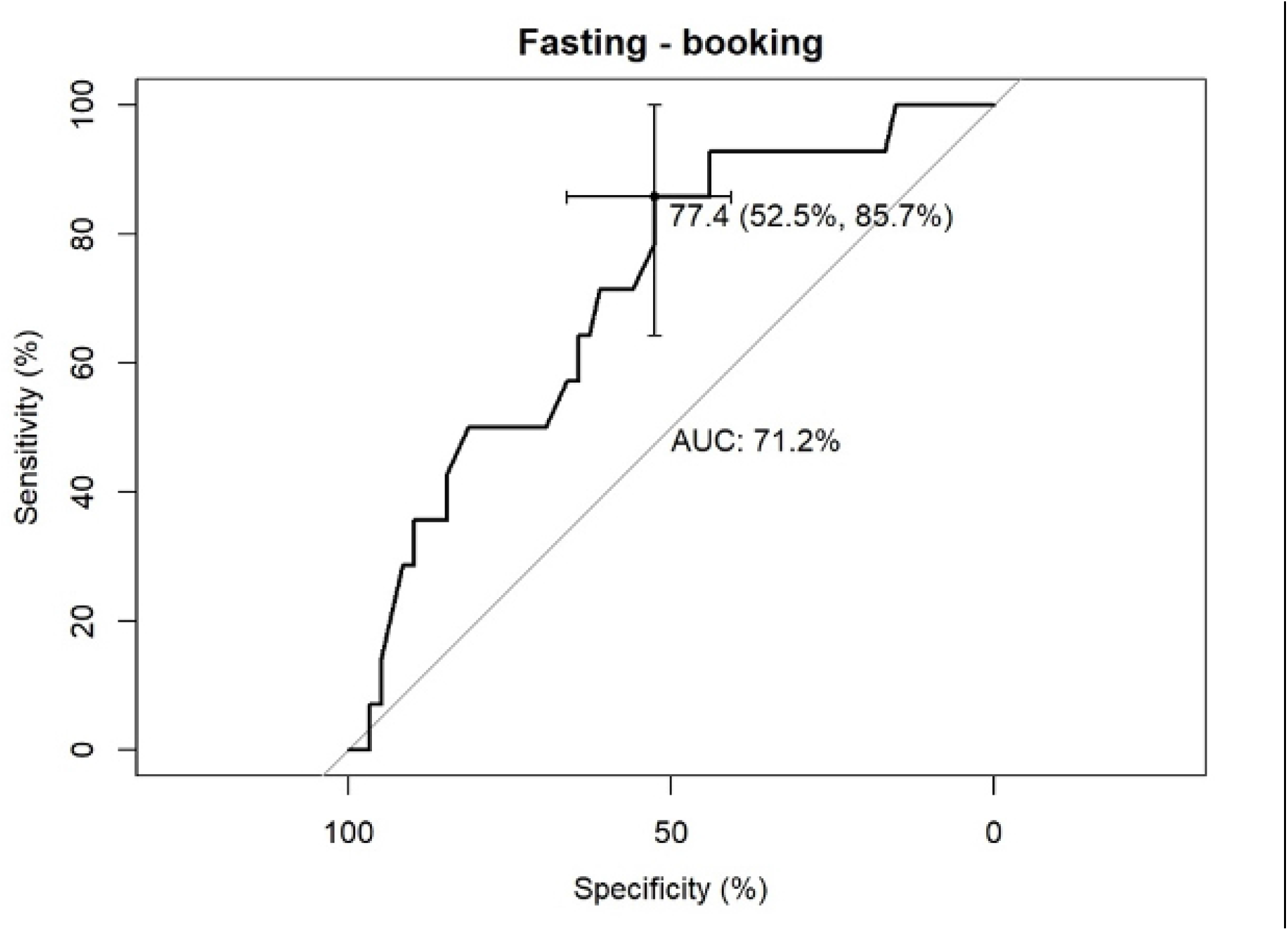
ROC curve for booking visit fasting blood glucose level

**Fig 8.**
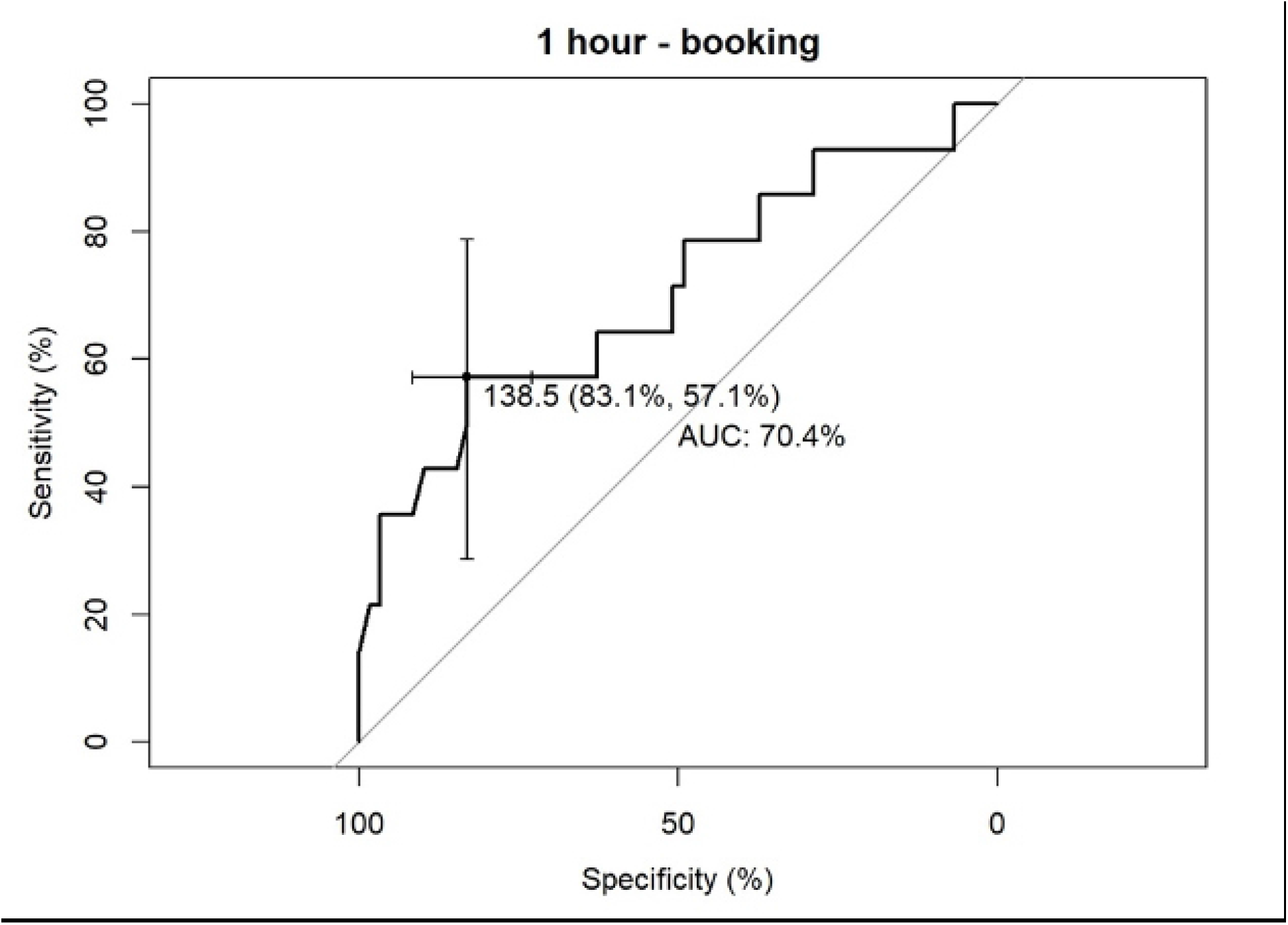
ROC curve for booking visit one-hour blood glucose level

**Fig 9.**
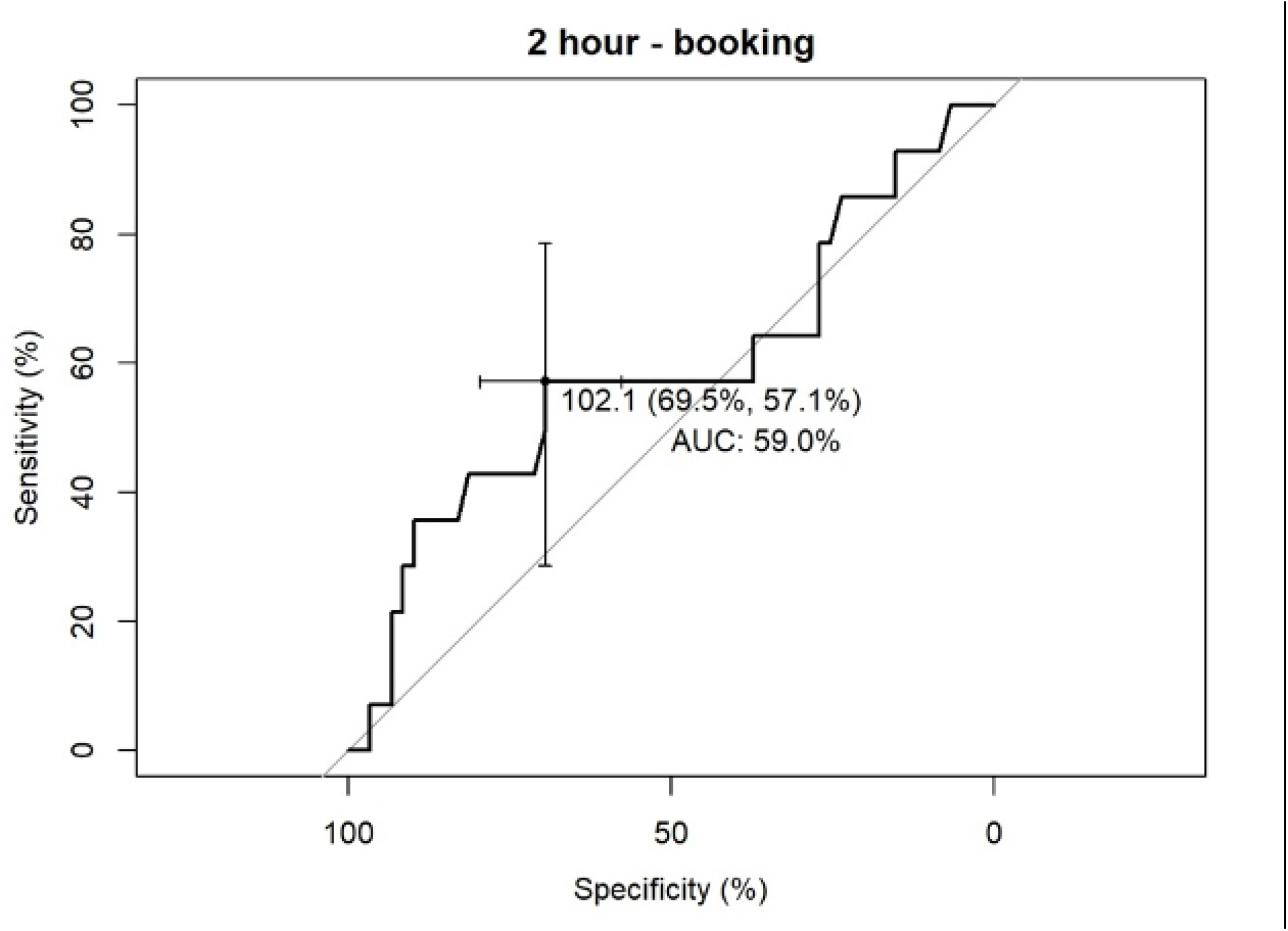
ROC curve for booking visit two-hour blood glucose level

The discriminative powers when booking visit plasma glucose values are combined together are summarized in Table 02.

**Table 02.** Discriminatory powers of different combinations of booking visit plasma glucose values

| Variables | Sensitivity (%) | Specificity (%) | AUC (%) |
| --- | --- | --- | --- |
| Fasting, one-hour and two-hour values | 85.7 | 67.8 | 79.3 |
| Fasting and one-hour values | 92.9 | 67.8 | 79.7 |
| Fasting and two-hour values | 85.7 | 57.6 | 71.3 |
| One-hour and two-hour values | 50.0 | 93.2 | 71.9 |

It can be noted that the combined booking plasma glucose values have a better discriminatory power when compared to that of isolated plasma glucose values. The ROC curves of fasting and one-hour combination, fasting and two-hour combination and the combination of fasting, one hour and two-hour plasma glucose values are depicted in Fig 10, Fig 11 and Fig 12 respectively.

**Fig 10.**
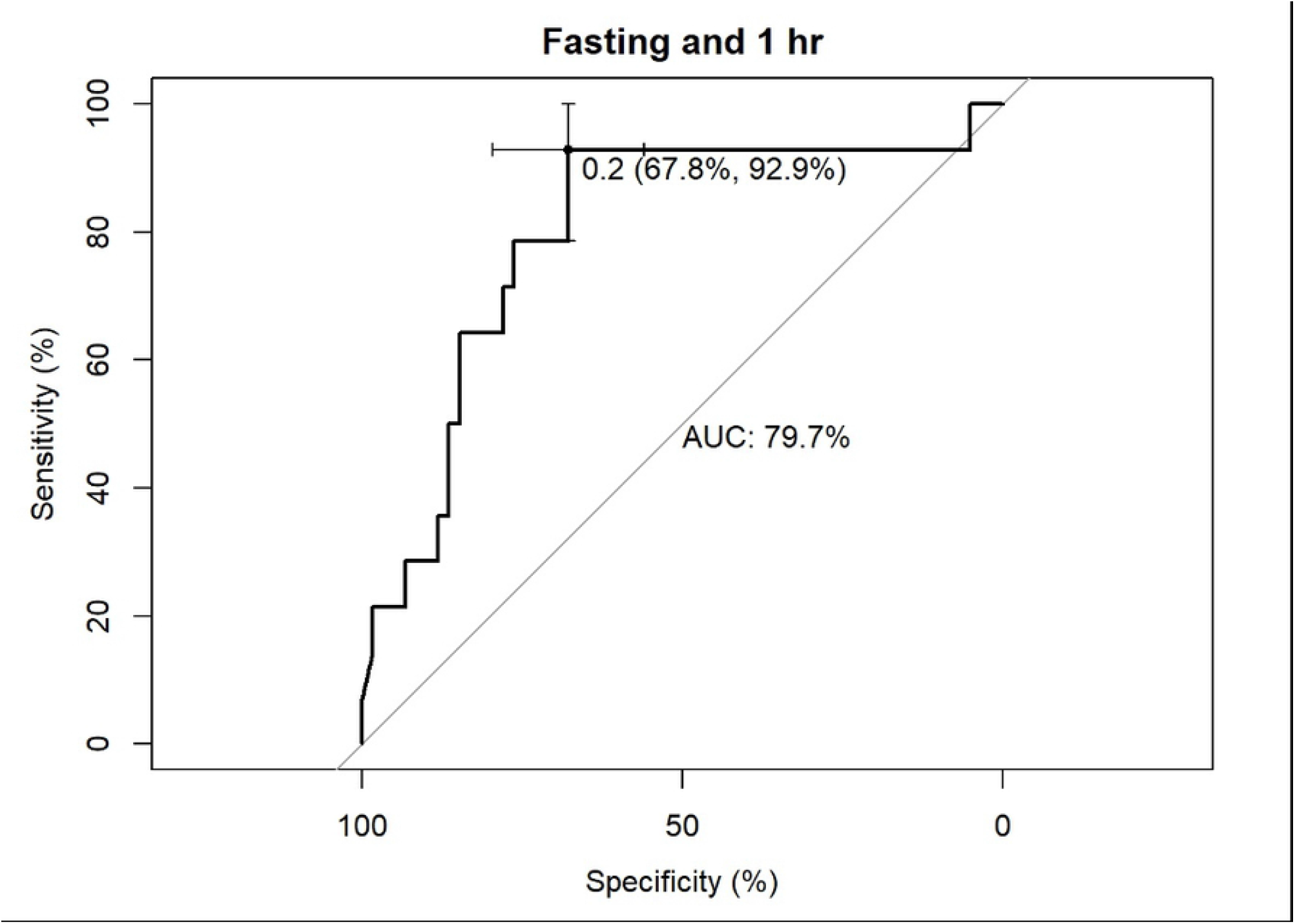
ROC curve for fasting and one hour plasma glucose combination

**Fig 11.**
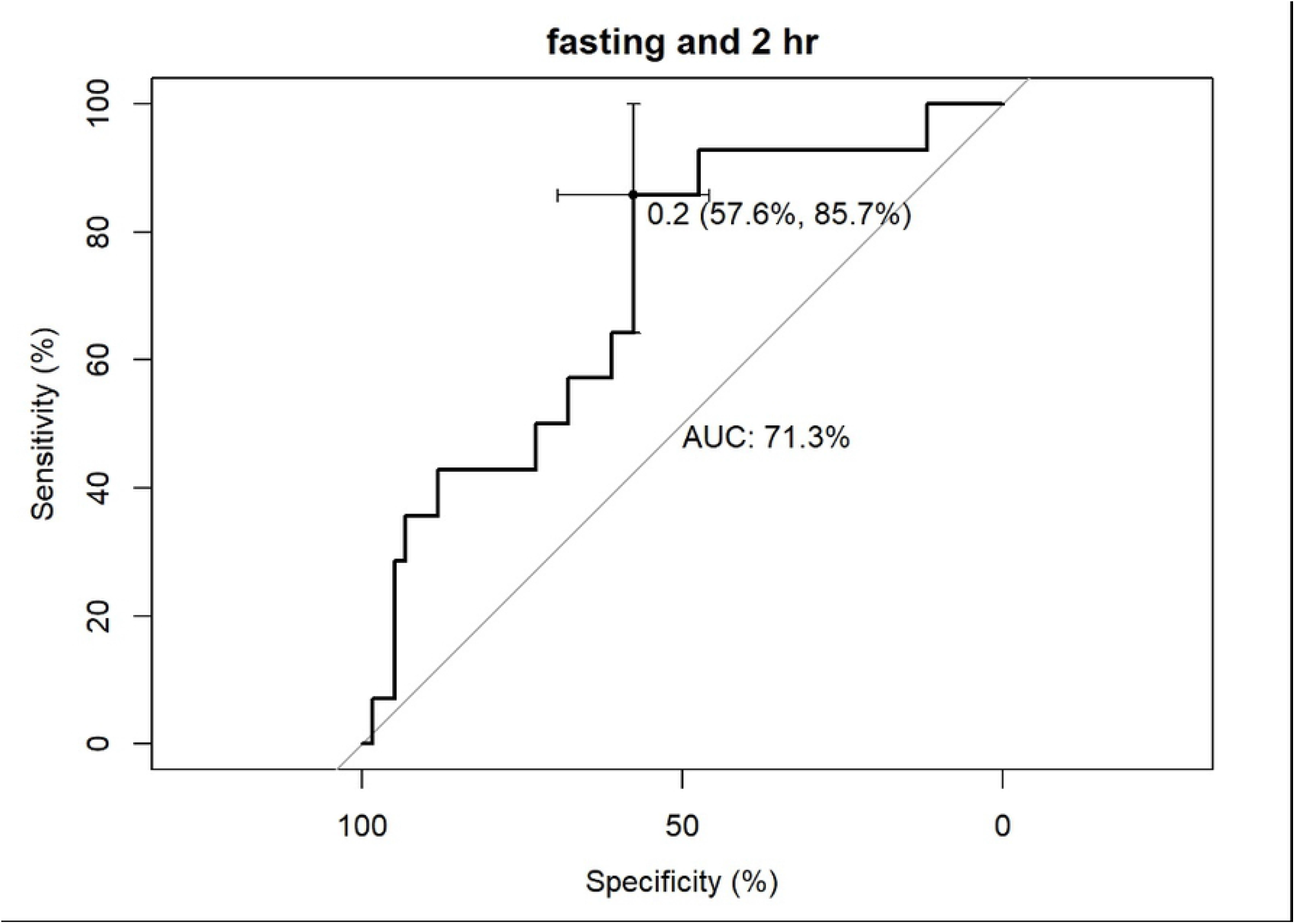
ROC curve for fasting and two hour plasma glucose combination

**Fig 12.**
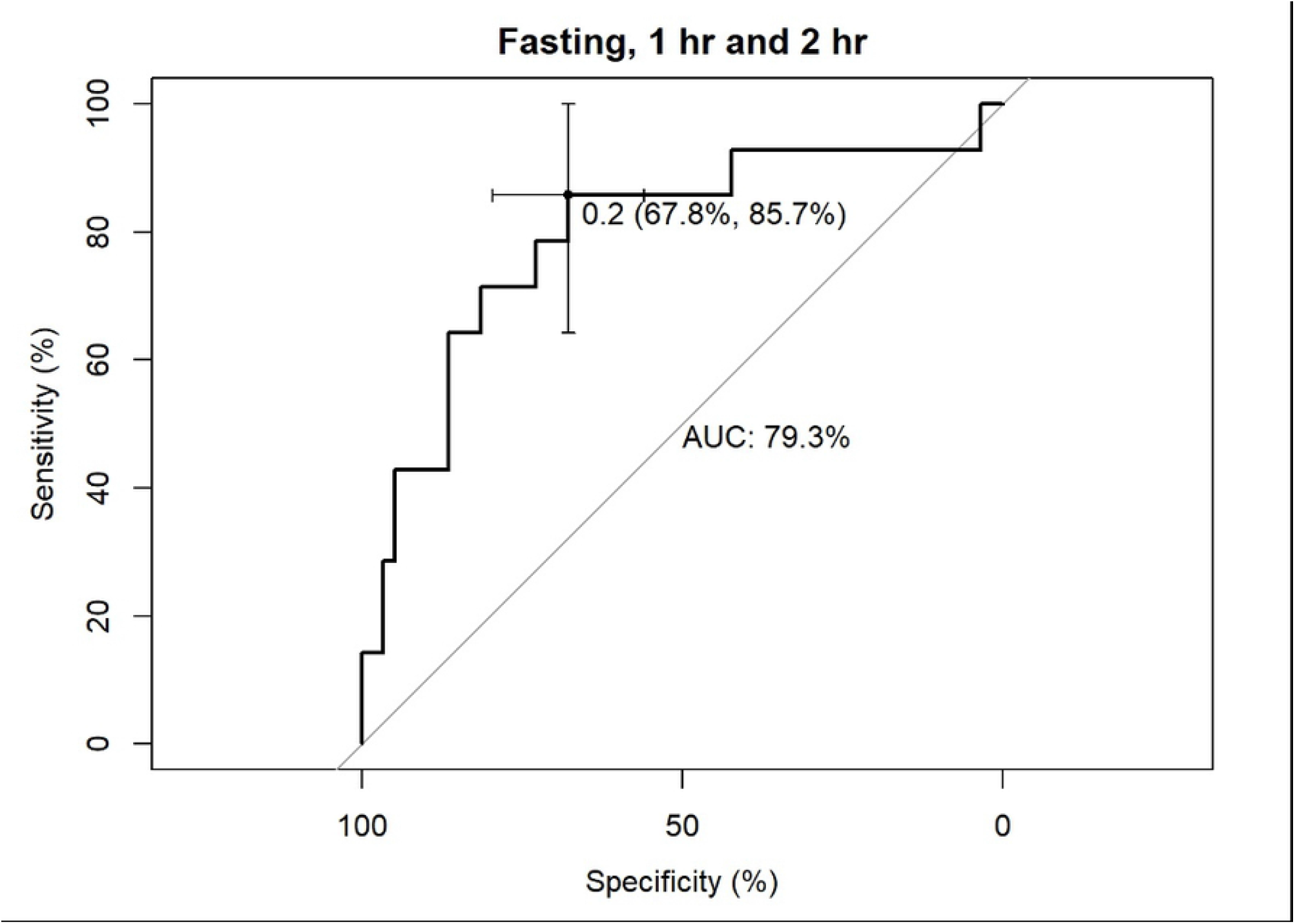
ROC curve for fasting, one hour and two hour plasma glucose combination

## Discussion

In Sri Lanka, all the mothers are offered screening for GDM as the population falls under a high-risk group due to South Asian ethnicity and hence undergo OGTT at 24-28 weeks. However, A narrative review published y Raets et al., indicate that mothers with early GDM are at more risk of adverse pregnancy outcomes. It further highlights that some pregnant women have hyperglycemia under the range of classification as diabetes at booking visit who will later be diagnosed with GDM at 24-28 weeks [6].

According to our study the individual values of fasting, one-hour and two-hour blood glucose individually does not predict the risk of future GDM with great accuracy. However, with regard to the combination models it can be noted that the model utilizing the fasting and one-hour plasma glucose values in the booking visit as input parameters was able to achieve an AUC of 79.7% for the ROC curve with a sensitivity of 92.86%, specificity of 67.80%, positive predictive value of 40.62% and a negative predictive value of 97.56%. Hence, in a woman with a normal OGTT this model is able to predict the chance of developing GDM in the future thus allowing preemptive lifestyle modification and more stringent surveillance.

Lekva et al. recruited 1031 pregnant women with the aim of identifying the use of OGTT at 14-16 weeks for the diagnosis of GDM by adjusting the cut offs. This study examined different cutoffs of early OGTT values for the prediction of GDM and found that approximately 80% sensitivity could be achieved when lowering fasting glucose cutoff by 8% and 60/120 minutes glucose cutoffs by 32% for IADPSG 2010 and WHO 2013 criteria. However, it concluded overall that early OGTT was not suitable to predict the occurrence of GDM [7].

In contrast, the logistic regression model proposed in this paper which takes into account both fasting and 1 hour blood glucose value, has the potential to predict the future occurrence of GDM with high sensitivity. This model can be utilized in antenatal care to identify women with a seemingly normal OGTT at the booking visit but with a high risk of developing GDM, enabling early lifestyle modifications and regular monitoring of blood glucose.

## Conclusion

A logistic regression model with fasting and one-hour plasma glucose values in the booking visit OGTT as input parameters, identifies women with the potential to develop GDM with high sensitivity.

## Data Availability

Data will be held in a repository after publication.

## Acknowledgements

We wish to acknowledge the staff of the antenatal clinics in Kandy for their support during the data collection process.

## Notes

### Competing Interest Statement

The authors have declared no competing interest.

### Funding Statement

The author(s) received no specific funding for this work

### Author Declarations

Ethics review committee, Faculty of Medicine, University of Peradeniya

